# Mechanisms of action of a community-led HIV prevention and treatment programme for female sex workers in Zimbabwe: a mixed-method analysis

**DOI:** 10.64898/2026.10.08.26365041

**Authors:** Joanna Busza, Harriet S Jones, Primrose Matambanadzo, Tsitsi Hove, Sungai T Chabata, Tatenda Kujeke, Memory Makamba, Gracious Madimutsa, Fortunate Machingura, Frances M Cowan, James R Hargreaves

## Abstract

**Background:** Community-led interventions for female sex workers (FSW) can improve HIV prevention and treatment outcomes by lowering self-stigma and strengthening social cohesion to improve service uptake. In Zimbabwe, the AMETHIST intervention trial integrated peer-led microplanning with self-help groups for FSW and demonstrated improved HIV treatment outcomes. We investigated the role of social cohesion and self-stigma in explaining these findings.

**Method:** We conducted a mixed-method analysis of data collected during AMETHIST. We re-analysed qualitative data from interviews and group discussions to identify narratives linking self-stigma, engagement with microplanners and/or self-help groups and HIV service use. We analysed quantitative data from endline respondent-driven sampling surveys for differences in self-reported measures of social cohesion and self-stigma between trial arms, and between those who did and did not participate in intervention activities or access services.

**Results:** In both arms, FSW self-reported engaging with peer supporters, participating in self-help groups and accessing HIV services. Qualitative narratives suggested peer support and group participation could reduce self-stigma and support service uptake. Lower levels of self-stigma were seen in the intervention arm (58.6% vs 63.2%; aOR 0.77; 95% CI 0.61-0.86). Social cohesion exceeded 50% across the whole sample, but was not associated with study arm, nor with engagement in intervention-related activities.

**Discussion:** Although qualitative data suggested some FSW felt their uptake of services was influenced by intervention components, quantitative analyses did not suggest either variable was on the causal pathway to the outcomes of the intervention. A limitation may have been the measurement of these complex constructs and the high levels of social cohesion across both study arms.

**Contributions to the literature:**

- Community-led, empowerment-based interventions are the cornerstone of effective HIV responses for sex workers. The hypothesised mechanisms of improved social cohesion and self-efficacy through which these work to increase HIV services uptake are less well understood.
- Our study contributes to evidence on the social mechanisms through which community based interventions work to improve HIV outcomes among female sex workers in Zimbabwe
- Our study found that female sex workers who more frequently accessed HIV services were more likely to report higher levels of social cohesion, but was unable to establish whether this was a mechanism along the pathway between intervention engagement and improved HIV outcomes Our study indicated that reductions in self-stigma through peer-led community-based support are important for HIV programme implementation; increasing the uptake of HIV services and improving HIV outcomes

## Background

Disproportionately high HIV prevalence among sex workers is well documented; global estimates in 2022 indicated a relative risk approximately nine times higher than that of the general population (1). Sex workers’ heightened vulnerability is driven by overlapping social and structural factors including criminalisation of sex work, stigma and discrimination, exposure to violence, and limited access to health services, all of which constrain prevention, testing, and treatment uptake (2–4). A wealth of literature (5–9), as well as HIV programming guidance from UNAIDS and the World Health Organization (WHO), recommends community-led, empowerment-based interventions as a cornerstone of effective HIV responses for sex workers (10–12). Such approaches position sex workers as leaders in programme design and implementation, requiring a strong foundation of community empowerment translated into collective action (13–15).

A key theoretical underpinning of these strategies is the concept of social cohesion, referring to the degree of mutual trust, solidarity, and shared identity among members of a community. In public health research, greater social cohesion has been associated with capacity to mobilise resources and advocate for rights and services (16, 17). Among female sex workers (FSW), social cohesion can increase the ability to negotiate condom use, respond collectively to violence, and challenge discriminatory practices (18–21). Social cohesion also strengthens social support networks through which sex workers can access emotional, financial, instrumental and other support, further enhancing resilience in the face of structural vulnerabilities (22–24). Associated health outcomes include reductions in STI and HIV prevalence and violence, and increases in ART uptake and adherence (25–27). The success of large scale programmes such as the Sonagachi Project and Avahan Initiative in India set the precedent for purposefully building social cohesion and social support as precursors to collective action (19, 28–30). Common strategies include peer outreach, “safe spaces” for group discussions, and participatory activities to bring FSW together to identify and act on shared concerns (31–34).

Although less common in sub-Saharan Africa, HIV programmes combining biomedical interventions such as condom promotion, HIV testing with referral for ART and PrEP, and syndromic management of STI with community mobilisation activities designed to increase social cohesion and support have been tailored for FSW (14, 35). However, initiating these processes can be challenging, particularly in contexts where FSW are marginalised, criminalised, and divided by competition and mistrust (36). The heterogeneity of FSW populations further shapes risk behaviours, peer support, and service engagement. Effective interventions thus need to both tailor support to individuals’ specific needs at the same time as they attempt to foster solidarity.

In Zimbabwe, the AMETHIST (Adapted Microplanning to Eliminate Transmissible HIV In Sex Transactions) intervention was designed with this dual focus, integrating two complementary community-led strategies: microplanning and self-help groups (SHG) (37). Microplanning applies a systematic, data-driven approach to peer outreach to ensure more vulnerable individuals receive more intensive engagement (38). Furthermore, by linking sex workers together and facilitating referrals to a wide range of services and resources (e.g. legal support), peer microplanners contribute to increasing social cohesion in the community (39). In parallel, SHG provide a structured platform for community-building and empowerment through formation of small, problem-solving groups. SHG have been shown to provide physical and psychosocial space for FSW to recognise shared experiences and concerns and move toward collective action for social change (32). In relation to HIV prevention and treatment, groups generate supportive relationships among members that can help reduce self-stigma, leading to improved self-efficacy to engage proactively with care (40, 41).

By combining microplanning’s targeted outreach with SHG’s focus on solidarity and joint action, the AMETHIST intervention aimed to reduce sex workers’ self-stigma and increase both individual self-efficacy and collective capacity to overcome barriers to HIV prevention and treatment, ultimately reducing HIV acquisition and transmission (Figure 1).

**Figure 1:**
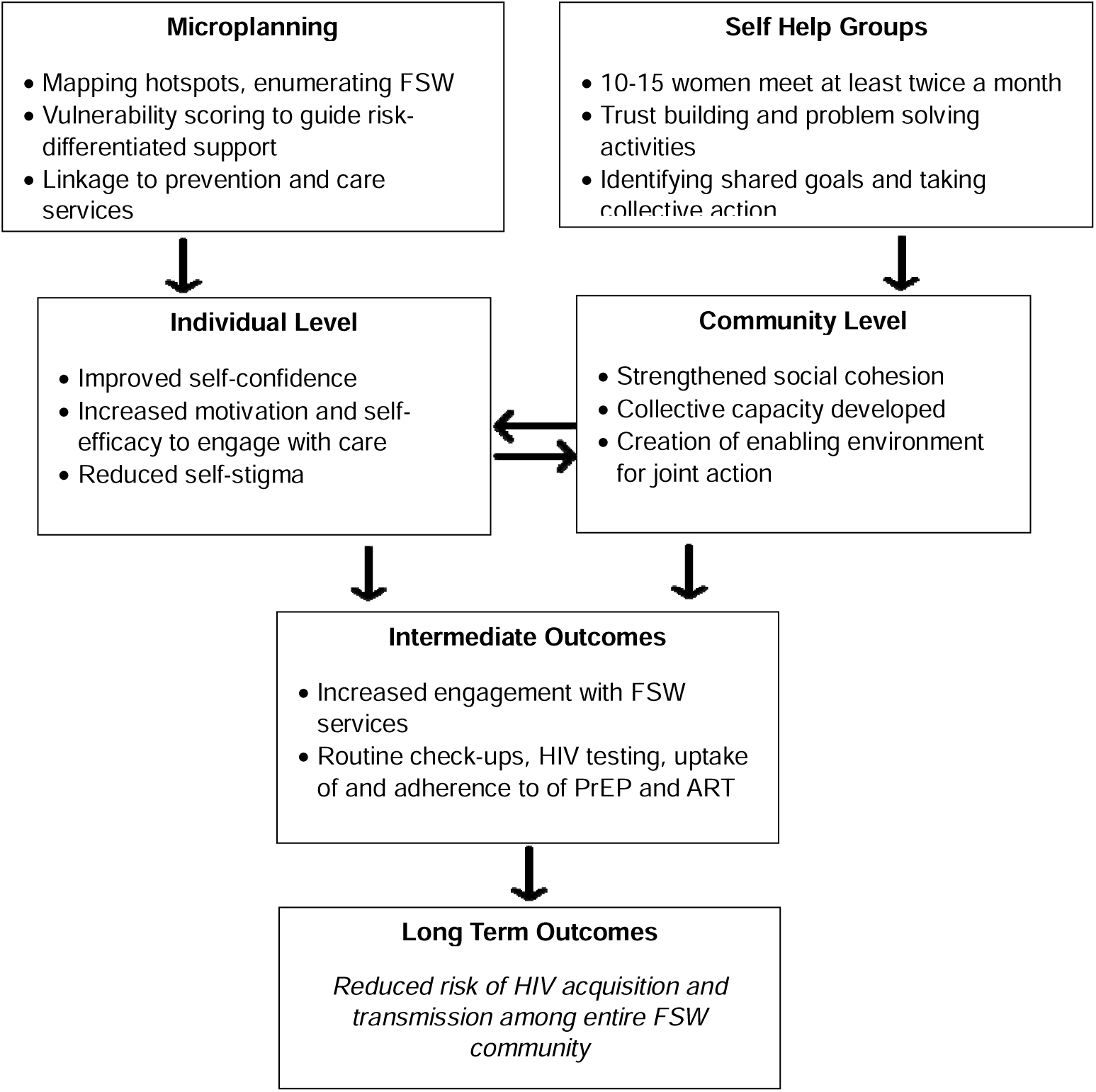
Flow diagram of the AMETHIST intervention change pathways.

We hypothesised that the AMETHIST intervention would influence HIV service uptake through two pathways. First, a psychosocial pathway, whereby engagement with the intervention would reduce internalised self-stigma and enhance self-efficacy, leading to increased service utilisation; and second a collective pathway, whereby participation in SHG would strengthen social cohesion, facilitating mutual support and collective capacity to engage with care

A cluster randomised trial conducted in 22 sites compared AMETHIST to the standard care offered by the national Key Population (KP) programme, but found no statistically significant difference between arms in the composite primary outcome of risk of HIV acquisition (in HIV negative women) or HIV transmission (in women living with HIV) (42). A stratified analysis found that among the HIV-positive women viral suppression rates were higher in the intervention arm, whereas consistent prevention behaviours such as condom use and PrEP adherence were not impacted by the intervention. However, uptake of HIV services was higher in intervention sites including clinic registrations, HIV tests and new diagnoses, and initiations of PrEP (42).

In this paper, we seek to explore whether the AMETHIST intervention achieved the observed increases in FSW use of HIV services through our hypothesised pathway of strengthening social cohesion and reducing self-stigma. We present both quantitative and qualitative data to understand relationships between microplanning and SHG and processes of building trust, reducing stigma, and willingness to work together among sex workers, and how these, in turn, influenced uptake of HIV prevention and treatment services.

## Methods

### Summary of Intervention

Described in detail and reported in line with the TIDieR checklist elsewhere (37, 43), the AMETHIST intervention trial (June 2019-December 2022) adapted microplanning to the Zimbabwean context, adding SHGs as an additional component. Twenty two clusters were purposively selected from 57 KP programme sites and restricted randomisation used to assign each cluster to intervention or control (42). FSW trained as microplanners mapped “hotspots” with high concentrations of sex work, estimated local FSW populations, and managed caseloads of 50–80 women. Mapping and population estimates were repeated every 8 months. Microplanners conducted outreach, assessed individual risk, and provided support tailored to each woman’s vulnerability, routinely tracking changes in FSW’s individual risk to determine the content and frequency of contact. Programme outreach workers debriefed and supervised microplanners weekly. Over 7000 FSW were microplanned in the 11 intervention sites.

Microplanners were also expected to establish SHGs of 10–15 FSWs who met fortnightly for self-directed discussions and activities. Each microplanner was meant to form at least two SHGs and reach 30–40% of their caseload, although due to restrictions on in-person meetings during the COV-SARS2 pandemic, most microplanners were able to start just one SHG. SHGs addressed a range of issues, including business or savings opportunities, childcare, personal hygiene and health, including encouraging one another to use condoms but also to attend recommended quarterly clinic visits for check-ups, HIV testing and follow-up initiation and adherence to PrEP or ART. In total, 65 SHG were established, with programme monitoring data showing that 30% of FSW registered with a microplanner were enrolled and 30 groups were still active at the end of the intervention period. In control sites, an existing model of clinics supported by peer educators continued. Peer educators did not systematically enumerate FSW in their catchment area, track a specified caseload of FSW, offer risk-differentiated levels of support, or organise SHG. While SHG were not offered by peer educators in control sites, women in Zimbabwe often join informal groups organised around pooling resources or saving money, known as mukando.

### Quantitative Data Collection and Analysis

A respondent-driven sampling (RDS) survey was conducted at the end of the AMETHIST trial for the outcome evaluation among 4444 FSW in the 11 intervention and 11 control sites. The survey was conducted between 18 October and 13 December 2021. For survey recruitment we used geographical and social mapping to identify six FSW ‘seeds’ in each cluster. Each was given two coupons to distribute to peers, who, once interviewed would receive their own coupons for referring two further peers until at least 200 women were recruited per site. Participants received $5 and an additional $2 for each woman they recruited. Eligible women had exchanged sex for money in the past 30 days, were aged 18 years or older, and had been living or working for at least 1 month in the trial site. Surveys were self-administered using an audio-computer assisted survey instrument (ACASI). Our endline survey methods are described in further detail elsewhere (42).

We analysed RDS survey data from the 22 clusters, excluding seed participants. Our analysis was RDS-II weighted to account for participants with larger social networks having a greater chance of being recruited. We first describe the population of surveyed FSW, by intervention and non-intervention study arms, by sociodemographic characteristics (i.e. age, marital status and education); sex work specific characteristics (i.e. age at first selling sex, sex work related income and breaks in sex work); their engagement in trial-related or KP programme activities (i.e. ever engagement with a SHG, contact with AMETHIST microplanners or peer educators); and HIV service use (i.e. CeSHHAR clinic attendance).

We generated an individual level measure of social cohesion based on a FSW’s belief in the capacity of sex workers to work together for mutual benefit. In our RDS survey we measured social cohesion across five questions: My colleagues will support me if I refuse to have sex with a client without a condom, my colleagues will help me if they see a client become aggressive or violent, I feel comfortable talking to other sex workers about work issues, I feel comfortable talking to other sex workers about health-related issues and Sex workers can improve their working conditions by working together). Each question had had five possible response options; strongly agree; agree; disagree; strongly disagree; no FSW colleagues, each denoted by a numeric score from 0 representing ‘no FSW colleagues’ to 4 representing ‘strongly agree’. We used Cronbach’s Alpha to assess the extent to which these items measured the same concept. We calculated an individual level score by summing scores across the five social cohesion items, with higher scores denoting greater social cohesion. We plotted the distribution of total social cohesion scores and used the median score across all five items as a cut off to generate a binary social cohesion variable for further analysis.

We followed the same process for measures of sex work-related self-stigma as a proxy measure for the extent to which FSW overcame internalised feelings of worthlessness and transitioned toward motivation and self-efficacy to make positive change, particularly regarding their health. Our survey did not directly measure self-efficacy but we have previously found high levels of internalised stigma related to sex work undermines FSW confidence in seeking services (44). We measured self-stigma through three questions on loss of respect or standing in the community, thinking less of oneself and feeling ashamed. Here we used three survey questions with five possible response options; strongly agree; agree; disagree; strongly disagree; I don’t know. We excluded responses of ‘I don’t know’ from our combined stigma measure after initially summarising our findings, with higher scores on our final variable denoting greater self-stigma.

As our survey was population-based it was not expected that all survey respondents would have engaged in the AMETHIST intervention or control KP programme, but that there would be increased social cohesion and reduced self-stigma in intervention compared to control sites. To understand whether the AMETHIST intervention strengthened social cohesion and reduced self-stigma we independently assessed the association of our binary measure of social cohesion and then self-stigma with study arm. We used logistic regression, with robust standard errors to account for clustering by study site, presenting crude OR and 95% CIs and then our estimates adjusted for age, HIV-status and sex work intensity as potential confounders identified a priori.

Pooling our data across study arms, we then explored differences in social cohesion and self-stigma by self-reported engagement with HIV services. We assessed crude and adjusted associations between our outcome variables and engagement with SHGs or peer educators. The type of SHG membership and engagement with a microplanner or peer educator was expected to be different between intervention and control arms, although captured under the same measure in our survey. For SHG membership we asked: Have you ever been part of any self-help group with other female sex workers in your community? For engagement with a microplanner (locally referred to as Empowerment Workers) we asked: Have you ever spoken to a Sisters/CeSHHAR Peer Educator or Empowerment worker? To investigate this further, we calculated stratified proportions and assessed effect modification in our adjusted logistic regression models by separately including SHG membership as an interaction term and then engagement with an empowerment worker to identify whether associations were different in the study arms.

We lastly investigated the association between our social cohesion and stigma measures and our intermediate outcome of increased FSW clinic engagement. We asked survey respondents whether: *In the past 12 months, how often have you attended the Sisters with a Voice clinic/CeSHHAR clinic?* We categorised responses into four or more times and fewer than four times. Using this binary variable as our outcome we first analysed the association between clinic attendance and social cohesion and then stigma, and then whether including clinic attendance as an interaction term in our models would identify any differences in association between intervention and control sites in our study. All our quantitative analyses were conducted in StataNow/SE v19.5.

### Qualitative Data Collection and Analysis

As part of a mixed-methods process evaluation embedded within the AMETHIST trial, we collected in-depth qualitative data in 3 of the 11 intervention sites, purposively selected to capture variation in region, sex work settings, population size, and programme scale. Each was visited within six months of intervention launch and again after 12–18 months to examine how microplanning and SHG activities were received and shaped by local context. The sites included a large town with university and mining activity (Chinhoyi, Northern Region), a smaller town near a military base with street- and venue-based sex work (Rusape, Eastern Region), and a rural truck stop on the South Africa highway (Ngundu, Southern Region).

At each site, semi-structured interviews were carried out with 1 outreach worker, 5 microplanners, and 5 FSW (including active participants, drop-outs, and non-participants) at both time points. Natural group discussions were held with 4 SHG members in each site, also purposively selected for diversity of experience (well-functioning, poorly functioning or disbanded). Trained qualitative fieldworkers conducted all interviews and group discussions in Shona at clinics, SHG meeting spaces, or, during COVID-19 restrictions, in outdoor locations where privacy could be assured. Sessions were recorded, transcribed into English, anonymised, and managed in NVivo for analysis.

For this paper, we interrogated the data deductively to identify whether and how microplanning and SHG influenced sex workers’ willingness and capacity to use health services. Analysis applied a priori codes anchored in the intervention’s key components: microplanning processes, SHG dynamics, and health service utilisation and our conceptual focus on social cohesion and self-stigma and their effects on self-efficacy.

### Ethical Approvals

The following ethical review boards approved the study: Medical Research Council of Zimbabwe (MRCZ/A/2559), the London School of Hygiene and Tropical Medicine (ref:19123), and the Liverpool School of Tropical Medicine (Ref 19-115RS). The AMETHIST intervention trial was conducted line with the Ottawa Statement on the Ethical Design and Conduct of Cluster Randomized Trials. Written informed consent from participants was obtained before enrolment.

## Results

### FSW characteristics, intervention and service engagement

After excluding seeds (n=132), 4,312 FSW were included in our RDS survey analysis. The median age of respondents was 31 (IQR 25-38) and similar in intervention sites at 30 (IQR 24-37). One fifth of all respondents, 20.5% (886/4312), reported that they had a period of at least three months in the previous year when they had not sold sex. This was higher in control sites, 23.0% (497/2158). Overall, 45.9% (1977/4311) of respondents tested HIV-positive, with higher HIV prevalence in control sites at 48.4% (1044/2158).

Across both trial arms, a quarter of respondents reported ever being part of a SHG with other FSW in their community (24.5%; 1,055/4312). Of these, 83.1% (881/1055) reported that their SHG group was still active. Engagement in a SHG was higher in the intervention arm at 27.6% (594/2154) compared to the control arm 21.4% (461/2158; OR 1.30; 95% CI 1.10-1.54), as was the likelihood of a SHG still being active (386/461, 83.7% vs 495/594, 83.3%; OR 1.30, 95% CI 1.08-1.55). Overall 57.4% (2,477/4312) of FSW reported having ever spoken to a peer educator or microplanner, with a similar proportion in each trial arm (Intervention: 59.1%, 1,272/2,154 vs Control: 55.8%, 1,205/2158; OR 1.11 95% 0.96-1.29). Overall 2,935/4312 FSW reported having ever attended a CeSHHAR programme clinic, with 30.5% 1,317/4,312 attending at least quarterly. Quarterly clinic attendance was greater in the intervention sites at 33.7% (727/2,154) compared to 27.3% (590/2,158) in control sites (OR 1.32, 95% CI 1.13-1.55) (Table 1).

**Table 1:** Characteristics of FSW overall and by study arm (N=4,312)

**Table 2:** Measures of social cohesion and self-stigma (N=4,312)

Our qualitative data suggest that microplanning could be an important means by which some FSW learned about the dedicated FSW clinics, and were more likely to consider attending because their peers provided regular and repeated endorsement

> *I first met [microplanner] in 2019 and she was the one who told me that there was something called Sisters’ Clinic [Rusape, FSW]*
>
> *There are some people who don’t know about the Sisters Clinic. Then there are some people who don’t even like to visit the clinic. If I talk to these people they might not listen to me but if you [a microplanner] talk to them then they are likely to listen. [Ngundu, FSW]*
>
> *She [microplanner] … just encourages us to visit the clinic. She can just be like, “where are you?”, like when I was in Mberengwa or wherever even in Chiredzi, “please go to the clinic”. [Ngundu, FSW]*

Microplanners’ use of risk scoring to determine frequency of contact helped ensure FSW most likely to be hesitant about clinical services received the most intensive encouragement.

> *If she [microplanner] doesn’t see me for a week or two at the clinic, she will come and visit me and find out why I have not been to the clinic in two weeks and asks me to show her the condoms I have with me [to check my supply]. [Chinhoyi, FSW]*

### Did Individual Self-Efficacy Increase?

AMETHIST intervention participants gave examples of how regular contact with microplanners and being a member of a SHG bolstered feelings of self-worth and increased motivation to address vulnerability to HIV and other health issues. Overcoming self-stigma seemed particularly salient in developing self-efficacy for care.

> *When you feel guilty you would call what you do “chihure” [being a whore] and not “sex work” and being involved in “chihure” is just not good. When the Sisters’ programme started, there were times when we could comfort each other here and there, and there were times when you could realise that your situation that you thought was tense was way better than someone else’s or that some of the things that I come across are not new; someone else could have encountered worse things or someone else could have encountered the same. So you would ask yourself how to handle your situation and we then shared a lot of things. For me, the programme comforted me [Ngundu, FSW]*
>
> *Yes, [we share] personal advice, like if we encounter any problems or the condom bursts, we shouldn’t be ashamed to go and get treatment. That we should always go and get checked up at the clinic. Things like that. [Chinhoyi, FGD2]*

Furthermore, receiving psychological, financial and instrumental support from other FSW (whether a microplanner or fellow SHG members) was described in FSW’s narratives as on their pathway to service use.

> *It benefits me because sometimes I don’t always treat myself well and the microplanner is a reminder that someone out there cares for me. She is concerned about how I am living and if things are going well for me. Having someone follow up on me makes me feel better. [Rusape, FSW]*
>
> *B: We accompany each other, if one of us falls ill and we don’t really understand the illness, we take her to the clinic for testing.*
>
> *M: Yes, I once went with one to the clinic when she was ill and she got tested, she thanked me later because she is now fit and is taking medication. [Chinhoyi, FGD 4)*

Self-stigma items measured in our endline RDS survey showed inter-item correlation (Cronbach’s alpha 0.76). Summed items had a median score of 7 (IQR 6-9) (out of a possible score of between 3 and 15), with higher scores denoting higher levels of self-stigma. A total of 22 respondents reported they did not know whether they felt experiences of stigma across all stigma items and were excluded from further analysis.

Higher self-stigma was reported by 60.1% (2614/4290) of FSW, but at lower levels in intervention sites at 56.8% (1256/2143) compared to 63.2% (1358/2147) in control sites (OR 0.77; 95% CI 0.62-0.96). This association remained when adjusted for age, HIV-status and having taken a break from sex work greater than 3 months in the preceding 12 months (aOR 0.77 (0.61-0.96) 95% CI 1.13-1.52) (Table 3).

**Table 3:** Distribution of self-stigma and social cohesion by study arm (N=4,312)

Conversely, higher levels of self-stigma were associated with membership of a SHG, with 63.6% (665/1,046) of SHG members reporting self-stigma compared to 60.1% (1,949/3,244) of non-SHG members (aOR 1.31; 95% CI 1.10-1.54). This association was the same in both study arms, with no evidence that the effect of being part of a SHGs on self-stigma was different between intervention and control sites (p=0.94) (Table 4). Lower levels of self-stigma were associated with microplanner/peer educator engagement (59.4% 1,467/2,470 vs 63.0% 1,147/1,820; aOR 0.82 95% CI 0.70-0.97). This association was similar in both intervention and control arms, with no evidence of effect modification (p=0.60) (Table 4).

**Table 4:** Modification of the effect of SHG attendance and engagement with an empowerment worker on self-stigma and social cohesion by study arm.

Quarterly clinic attendance was less common among those reporting self-stigma, with 56.8% (745/1,317) of those attending four or more times a year reporting self-stigma, compared to 62.8% (1869/2,995) of those attending a clinic less than quarterly (aOR 0.75 95% CI 0.62-0.89) This association was also similar in intervention and control arms (p=0.50). (Table 5)

**Table 5:** Modification of the effect of self-stigma and social cohesion on quarterly clinic attendance by study arm.

### Did Social Cohesion Improve?

We found little evidence of a difference in social cohesion between arms across the FSW community, either from RDS survey or from the qualitative data. The SHG component of AMETHIST was specifically dedicated to creating an enabling environment for risk reduction and service use by building trust and cooperation between FSW to increase their knowledge and skills for collective action. Qualitative data point to mixed experiences regarding the extent to which FSW were able to get along and work for common good.

> *Looking at the year I started sex work and now, I would say that things have changed. When I started sex work, when I would go to the bars, people were in cliques, if fights broke out between the cliques, the fights would get so violent and nasty. Now that doesn’t happen as often. If a new FSW comes, we welcome her then we tell her about CeSHHAR and the next time we will be visiting the clinic and invite her to come with us. [Chinhoyi, FSW]*
>
> *Yes here in Ngundu things have improved, and it’s mainly because of CeSHHAR. Before CeSHHAR came with its programmes there were lots of fights, most people lost teeth and eyes fighting over men. But now it’s different we get to meet and share ideas. [Ngundu, FSW]*

At the same time, FSW noted the lack of harmonious relationships and the perseverance of competition and hostility.

> *Groups are difficult and are characterized by a lot of gossip. When you gather together as a group, you will be pretending that you are the same people but when you are not together, people gossip. [Ngundu, FSW]*
>
> *The problem these days is people are jealous, they are no longer as united as they used to be back then. Back then it was well known that no sex worker would get beaten up in the presence of other sex workers but these days other sex workers can cause the beating of another sex worker. So it’s now different from the past. [Rusape, FSW]*

Nonetheless, some SHG were effective in creating solidarity at the level of their membership, giving specific examples of collective capacity to solve both individual and shared problems. In the first case, members rallied around one of their peers following a medical emergency, while in the second, the SHG established a fund to deal with just such emergencies in future.

> *We contribute money, one example is of a lady who stays here, she gave birth to premature babies. We had to help and support her until she was discharged from the hospital. We looked after her. She is a sex worker who got pregnant and was abandoned by the father who is a married man. [Chinhoyi, FSW]*
>
> *What motivated the establishment of this group, we had one of us who fell sick and failed to go to work. That is when we came together and discussed that one of us is sick and failing to even go to the hospital. We then decided to form a group where each one of us will be contributing money in case one of us falls sick we can help each other. [Chinhoyi, FGD3]*

Many SHG set up traditional savings schemes (mukando) used to purchase groceries or household goods in bulk or set up funeral savings plans. However, our five RDS survey questions did not demonstrate an association between the intervention and higher social cohesion scores. Respondents scored a median of 22 (IQR 20-24) with a higher score (maximum 25) denoting higher levels of social cohesion, with a minimum possible score of 5 denoting low levels of social cohesion (Cronbach’s alpha 0.80). Social cohesion was high across both trial arms, with little room to improve on scores and high social cohesion represented in both our binary variable categories. We found no evidence of a difference in social cohesion between intervention and control sites (51.1% 1,100/2,154 vs 50.3%, 1086/2,158; OR 1.01 95% CI 0.78-1.32). This lack of an association remained after adjusting for age, HIV status and breaks in sex work (aOR 1.03 95% CI 0.79-1.33). (Table 2)

Adjusted for age, HIV status and sex work intensity social cohesion was higher among survey respondents who had engaged with either a peer educator or a microplanner; aOR 1.36; 95% CI: 1.10, 1.77. This association was similar in both trial arms with no evidence of interaction (p=0.64). There was no evidence that SHG membership was associated with greater social cohesion (aOR 1.06 95% CI 0.90-1.24) which was the same in both intervention and control arms. (Table 3, Table 4)

Quarterly clinic attendance was more likely among those reporting higher levels of social cohesion, with 57.4% (756/1317) of those reporting visiting a clinic 4 or more times in a year reporting high levels of social cohesion compared to 47.8% (1430/2995) of those who did not (aOR 1.44 95% CI 1.23-1.69). This association was similar in intervention and control sites. (Table 5)

## Discussion

In order to explore elements of our programme theory, we analysed qualitative and quantitative data to assess whether FSW exposed to the AMETHIST intervention package in Zimbabwe, and who engaged with peer-educators or participated in self-help groups, also reported higher levels of social cohesion and lower levels of self-stigma. Self-stigma appeared to be lower among FSW in intervention sites. The regular and tailored counselling visits by microplanners engendered feelings of being cared for and “mattering,” which have been documented as important contributors to self-confidence and self-efficacy, particularly among marginalised communities (44, 45). Self-stigma was also lower among those who had engaged with microplanners / peer educators, reflecting findings in other settings taking a community mobilisation approach (6, 46, 47).

We went on to assess whether social cohesion and self-stigma were associated with engagement with HIV prevention and treatment services. (43). Our quantitative analyses showed greater service use among FSW reporting higher levels of social cohesion, although with mixed findings in our qualitative data. Cross-sectional studies in other African contexts suggest that social cohesion is associated with positive HIV behavioural and health outcomes including higher condom use and lower reported social discrimination, including from the police (20). Among another key population, men who have sex with men, a study from Lesotho found an inverse association between an individual’s perception of their community’s social cohesion and depression (48). The importance of active participation has also been highlighted, often considered as an output of good social cohesion whereby community members who perceive trust and solidarity develop “collective capacity” to take concrete steps to further shared aims (20, 40).

Communities are not, of course, homogeneous in nature nor devoid of internal power dynamics and competition. Others have raised concern about simplistic characterisations of “communities” as though they are unified entities waiting to be “mobilised” (49); we have ourselves documented the very real barriers to creating shared community identity among FSW in Zimbabwe, although we found these not to be intractable given contextually-appropriate social investments and, critically, time for genuine trust to develop (49). It may be that the relatively small proportion (30%) of microplanned FSW who enrolled in SHG and the fact that only just over half SHG remained functional throughout the trial period was inadequate for shifting broader community norms. AMETHIST was delivered when Covid-19 restrictions were imposed on travel and gatherings and our SHG lost momentum during times when they were reliant on remote communication. This fundamentally undermined the core function of SHG, which were designed to bring FSW together to overcome distrust and competition and forge links of solidarity. Such processes are notoriously slow and often arduous, and unlikely to be amenable to remote communication (especially in a context of low phone ownership and ability to pay for data or airtime).

To what extent community mobilisation can be catalysed from outside a community has been a topic of longstanding debate; a recent systematic review highlighted the lack of clarity around whether social ties can be purposively altered by HIV interventions in ways that bring about positive change (31). Many authors have critiqued the lack of conceptual clarity around social cohesion and related concepts i.e. social capital, collective capacity, community empowerment (50), which has undoubtedly contributed to the diversity in operationalising measures for each of these (51).

Aggregating individual responses to survey questions has been criticised for not reflecting collective properties of a community (52, 53). One alternative that has been proposed is assessing social participation rates (54), akin to our measure of SHG participation. We found this to be similar between intervention and control arms, which would again suggest little difference in social cohesion. Nonetheless, it is likely that our measures for both social cohesion and self-stigma did not do justice to these complex ideas.

While our qualitative data provided more evidence of a pathway from SHG and peer support to motivation and self-efficacy to engage with health services, this partly reflects social desirability bias found in a setting where any attention to the needs of FSW goes much appreciated (55). We have previously shown that approval and acceptance of Zimbabwe’s national Key Population programme is high (56). We conducted natural group discussions with SHG members, who self-selected into them. Despite purposively sampling SHG with poor outcomes (including groups that collapsed), it is likely participants were particularly motivated to engage in collective activities, as found in a similar programme in India (57). Limitations in measuring SHG support through our quantitative analysis meant we were unable to identify whether FSW were engaging with CeSHHAR delivered services or services delivered by other providers. Specifically, AMETHIST SHG were not run in control sites, yet similar proportions in both arms reported having ever been part of a SHG with other sex workers. While we managed this at the analysis stage it is unlikely that we were able to fully assess the impact of SHG in this study.

A key strength of this study lies in its use of both quantitative and qualitative data from the AMETHIST trial, which enabled a comprehensive examination of one of the central hypotheses regarding the underlying mechanisms of change associated with our intervention. A related limitation, however, is the absence of quantitative measures that directly correspond to the theoretical constructs of interest, which constrains our ability to robustly quantify some of the proposed pathways of effect. Our analysis was limited to estimating associations between components of our proposed pathways due to the cross-sectional nature of our survey. We were unable to assess the temporal ordering of intervention exposure and self-stigma or social cohesion. Our quantitative measure of social cohesion was likely to be subject to a ceiling effect, with high levels of social cohesion reported among all survey respondents and making assessment of social cohesion difficult.

## Conclusion

Our analysis provides insights into the pathways through which the AMETHIST intervention may have influenced engagement with HIV prevention and treatment services among female sex workers in Zimbabwe. While we did not observe an effect on composite behavioural outcomes, evidence from secondary programme data and qualitative findings suggests that microplanning and SHG participation contributed to increased clinic attendance, HIV testing, and PrEP and ART initiations. Enhanced self-efficacy—manifested through reduced internalised stigma, feelings of being cared for, and increased confidence—emerged as a plausible mechanism underlying these improvements. Conversely, we found limited evidence that the intervention shifted broader measures of social cohesion, likely reflecting the relatively low SHG coverage, disruptions caused by Covid-19, and measurement limitations.

## Supporting information

Mechanisms of Action_TABLES_SUBMISSION

## Data Availability

All data produced in the present study are available upon reasonable request to the authors

## Funding declaration

The Key Populations (formerly Sisters’) Programme was funded through the Global Fund to fight AIDS, Tuberculosis and Malaria and the US President’s Emergency Plan for AIDS Relief through the US Agency of International Development. The AMETHIST intervention was funded by the Elton John AIDS Foundation. FMC and JRH were partly funded by the Wellcome Trust (214280/Z/18/Z). FMC and JRH were partly funded by the Bill & Melinda Gates Foundation (INV-007055).

## Data availability statement

The datasets analysed for the current study are available from the corresponding author on reasonable request. The request will be considered by the trial investigators and where agreed, prespecified data will be made available subject to a written proposal and a signed data sharing agreement.

## Notes

### Competing Interest Statement

The authors have declared no competing interest.

### Clinical Trial

The AMETHIST trial is registered with the Pan African Clinical Trials Registry, PACTR202007818077777

### Author Declarations

The following ethical review boards approved the study: Medical Research Council of Zimbabwe (MRCZ/A/2559), the London School of Hygiene and Tropical Medicine (ref:19123), and the Liverpool School of Tropical Medicine (Ref 19-115RS)

