## Supplementary material for "Mechanisms of action of a community-led HIV prevention and treatment programme for female sex workers in Zimbabwe: a mixed-method analysis": Mechanisms of Action_TABLES_SUBMISSION

| **Table 1: Characteristics of FSW overall and by study arm (N=4,312)** | | | |
| --- | --- | --- | --- |
| **Characteristics** | **TOTAL** | Control | **Intervention** |
|  | N=4,312  (RDS-II %) | N = 2158  (RDS-II %) | N=2,154  (RDS-II %) |
| **Demographics** |  |  |  |
| Age |  |  |  |
| Median (IQR) | 31 (25-38) | 32 (26-39) | 30 (24-37) |
| Education |  |  |  |
| None | 114 (2.7) | 76 (3.5) | 38 (1.8) |
| Primary | 983 (22.8) | 483 (22.4) | 500 (23.2) |
| Secondary | 3079 (71.5) | 1537 (71.4) | 1542 (71.6) |
| Tertiary | 131 (3.0) | 58 (2.7) | 73 (3.4) |
| *Missing* | *5* | *4* | *1* |
| Marital status |  |  |  |
| Never married | 1082 (25.1) | 445 (20.6) | 637 (29.6) |
| Married | 242 (5.6) | 114 (5.3) | 128 (5.9) |
| Divorced/separated | 2422 (56.2) | 1268 (58.8) | 1154 (53.6) |
| Widowed | 566 (13.1) | 331 (15.3) | 235 (10.9) |
| HIV status |  |  |  |
| Negative | 2334 (54.1) | 1114 (51.6) | 1220 (56.7) |
| Positive | 1977 (45.9) | 1044 (48.4) | 933 (43.3) |
| *Missing* | *1* | *0* | *1* |
| **Sex Work** |  |  |  |
| Do you consider yourself a sex worker? |  |  |  |
| No | 67 (1.6) | 33 (1.5) | 34 (1.6) |
| Yes | 4150 (96.2) | 2080 (96.4) | 2070 (96.1) |
| Sometimes | 95 (2.2) | 45 (2.1) | 50 (2.3) |
| Age first sold sex |  |  |  |
| Median (IQR) | 21 (18-26) | 22 (18-27) | 20 (18-25) |
| In the past year has there ever been a time when you did not sell sex for at least since 3 months? |  |  |  |
| No | 3426 (79.5) | 1661 (77.0) | 1765 (81.9) |
| Yes | 886 (20.5) | 497 (23.0) | 389 (18.1) |
| Income through sex work (past 3 months) |  |  |  |
| Very little, less than a quarter | 1522 (35.3) | 817 (37.9) | 705 (32.7) |
| Some, between a quarter and half | 1259 (29.2) | 631 (29.2) | 628 (29.2) |
| Most, more than half, but not all | 724 (16.8) | 316 (14.6) | 408 (18.9) |
| All, only source of income | 807 (18.7) | 394 (18.3) | 413 (19.2) |
| **Service engagement** |  |  |  |
| Have you ever been part of any self-help group with other female sex workers in your community |  |  |  |
| No | 3257 (75.5) | 1697 (78.6) | 1560 (72.4) |
| Yes | 1055 (24.5) | 461 (21.4) | 594 (27.6) |
| Is yes - is your SHG still active? |  |  |  |
| No | 174 (16.5) | 75 (16.3) | 99 (16.7) |
| Yes | 881 (83.5) | 386 (83.7) | 495 (83.3) |
| Have you ever spoken to a Sisters/CeSHHAR peer educator or microplanner? |  |  |  |
| No | 1835 (42.6) | 953 (44.2) | 882 (41.0) |
| Yes | 2477 (57.4) | 1205 (55.8) | 1272 (59.0) |
| In the past 3 months did you receive any support from a peer educator/ microplanner to attend the sisters clinic? |  |  |  |
| No | 1335 (31.0) | 683 (61.7) | 652 (30.3) |
| Yes | 2977 (69.0) | 1475 (68.3) | 1502 (69.7) |
| Have you ever attended the CeSHHAR Clinic/ Sisters with a Voice clinic? |  |  |  |
| No | 1377 (31.9) | 738 (34.2) | 639 (29.7) |
| Yes | 2935 (68.1) | 1420 (65.8) | 1515 (70.3) |
| In the past 12 months, Have you attended the Sisters with a Voice/CeSHHAR clinic? |  |  |  |
| No | 1377 (31.9) | 738 (34.2) | 639 (29.7) |
| Yes | 2935 (68.1) | 1420 (65.8) | 1515 (70.3) |
| In the past 12 months, how often have you attended the Sisters with a Voice clinic/CeSHHAR clinic? |  |  |  |
| Less than 4 times | 2995 (69.5) | 1568 (72.7) | 1427 (66.3) |
| 4 or more times | 1317 (30.5) | 590 (27.3) | 727 (33.7) |
| **Asked of FSW reporting that they had: Since you started being a sex worker has there ever been a period of 3 months or more when you did not sell sex ?* | | | |

| **Table 2: Measures of social cohesion and self-stigma (N=4,312)** | | | | | |  |  | |  | |  | |
| --- | --- | --- | --- | --- | --- | --- | --- | --- | --- | --- | --- | --- |
| **Social cohesion** | Strongly agree (5) | Agree (4) | Disagree (3) | | Strongly disagree (2) | | | No colleagues (1)*^$^* | | Median (IQR) | | Cronbach's Alpha |
| My colleagues will support me if I refuse to have sex with a client without a condom | 1843 (42.7) | 1427 (33.1) | 717 (16.6) | | 265 (6.2) | | | 60 (1.4) | | 4 (4-5) | |  |
| My colleagues will help me if they see a client become aggressive or violent | 2137 (49.6) | 1670 (38.7) | 319 (7.4) | | 113 (2.6) | | | 73 (1.7) | | 4 (4-5) | |  |
| I feel comfortable talking to other sex workers about work issues | 2176 (50.5) | 1827 (42.4) | 202 (4.7) | | 57 (1.3) | | | 50 (1.2) | | 4 (4-5) | |  |
| I feel comfortable talking to other sex workers about health-related issues | 2202 (51.1) | 1791 (41.5) | 223 (5.2) | | 47 (1.1) | | | 49 (1.1) | | 5 (4-5) | |  |
| Sex workers can improve their working conditions by working together | 1969 (45.7) | 1975 (45.8) | 258 (6.0) | | 68 (1.6) | | | 42 (1.0) | | 4 (4-5) | |  |
| TOTAL |  |  |  | |  | | |  | | 22 (20-24) | | 0.801 |
| **Self-stigma** | Strongly agree (4) | Agree (3) | Disagree (2) | | Strongly disagree (1) | | | I don't know (0) | | Median (IQR)* | | Cronbach's Alpha* |
| Because of my work as a sex worker, I have lost respect or standing in the community | 1184 (27.5) | 1366 (31.7) | 1117 (25.9) | | 340 (7.9) | | | 305 (7.1) | | 3 (2-4) | |  |
| Because of my work as a sex worker, I think less of myself | 801 (18.6) | 1069 (24.8) | 1767 (41.0) | | 594 (13.8) | | | 81 (1.9) | | 2 (2-3) | |  |
| Because of my work as a sex worker, I have felt ashamed | 730 (16.9) | 986 (22.9) | 1915 (44.4) | | 624 (14.5) | | | 57 (1.3) | | 2 (2-3) | |  |
| TOTAL |  |  |  | |  | | |  | | 7 (6-9) | | 0.757 |
| ** excludes respondents with 'I don't know' responses on all self-stigma questions (n=22)* | | | |  | |  |  | |  | |  | |
| *^$^ 7 FSW report ‘no colleagues’ for all questions* | | | |  |  |  |  |  |  |  |  |  |

| **Table 3: Distribution of self-stigma and social cohesion by study arm, self-help group membership and peer educator engagement (N=4,312)** | | | | | |  |
| --- | --- | --- | --- | --- | --- | --- |
|  | **TOTAL**  N (RDS-II %) | n (RDS-II %) | n (RDS-II %) | OR | aOR | p-value |
| **Study Arm** |  | Control | Intervention |  |  |  |
| **Self-stigma** | 4,290 | 2,147 | 2,143 |  |  |  |
| Low | 1676 (30.1) | 789 (36.8) | 887 (41.4) | 1 | 1 |  |
| High | 2614 (60.9) | 1358 (63.2) | 1256 (58.6) | 0.77 (0.62-0.96) | 0.77 (0.61-0.96) | 0.023 |
| **Social cohesion** | 4,312 | 2,158 | 2,154 |  |  |  |
| No | 2126 (49.3) | 1072 (49.7) | 1054 (48.9) | 1 | 1 |  |
| Yes | 2186 (50.7) | 1086 (50.3) | 1100 (51.1) | 1.01 (0.78-1.32) | 1.03 (0.79-1.33) | 0.850 |
| **Self-Help Group** |  | **No** | **Yes** |  |  |  |
| **Self-stigma** | 4,290 | 3,244 | 1,046 |  |  |  |
| Low | 1676 (30.1) | 1,295 (39.9) | 381 (36.4) | 1 | 1 |  |
| High | 2614 (60.9) | 1,949 (60.1) | 665 (63.6) | 1.27 (1.08-1.50) | 1.31 (1.10-1.54) | 0.004 |
| **Social cohesion** | 4,312 | 3,257 | 1,055 |  |  |  |
| No | 2126 (49.3) | 1,634 (50.2) | 492 (46.6) | 1 | 1 |  |
| Yes | 2186 (50.7) | 1,623 (49.8) | 563 (53.4) | 1.06 (0.90-1.25) | 1.06 (0.90-1.24) | 0.507 |
| **Microplanner/Peer Educator** | | **No** | **Yes** |  |  |  |
| **Self-stigma** | 4,290 | 1,820 | 2,470 |  |  |  |
| Low | 1676 (30.1) | 673 (37.0) | 1,003 (40.6) | 1 | 1 |  |
| High | 2614 (60.9) | 1,147 (63.0) | 1,467 (59.4) | 0.81 (0.69-0.95) | 0.82 (0.70-0.97) | 0.018 |
| **Social cohesion** | 4,312 | 1,835 | 2,477 |  |  |  |
| No | 2126 (49.3) | 1,006 (54.8) | 1,120 (45.2) | 1 | 1 |  |
| Yes | 2186 (50.7) | 829 (45.2) | 1,357 (54.8) | 1.38 (1.13-1.69) | 1.36 (1.10-1.67) | 0.004 |

| **Table 4: Modification of the effect of SHG attendance and engagement with an empowerment worker on self-stigma and social cohesion by study arm** | | | | | | | |
| --- | --- | --- | --- | --- | --- | --- | --- |
|  |  | **Low self-stigma** | | **High self-stigma** | | Adjusted* odds of self-stigma within intervention strata  (aOR; 95% CI) | **p-value** |
| Intervention status | **SHG / non-SHG (n=1046/3244)** | non-SHG (n=1295) | SHG (n=381) | Non-SHG (n=1949) | SHG (n=665) |  |  |
| **Control** | 455/1692 | 639 | 150 | 1,053 | 305 | 1.32 (1.02-1.69) | 0.03 |
| **Intervention** | 591/1552 | 656 | 231 | 896 | 360 | 1.30 (1.04-1.62) | 0.02 |
| *Measure of effect modification on the multiplicative scale: Ratio of OR 0.99 (0.71-1.38), p-value 0.94* | | | | | |  |  |
|  | **EW / non-EW**  **(n=1,835/2,477)** | EW (n=673) | non-EW (n=1003) | EW (n=1147) | Non-EW (n=1467) |  |  |
| **Control** | 953/1,205 | 336 | 453 | 610 | 748 | 0.87 (0.70-1.07) | 0.19 |
| **Intervention** | 882/1,272 | 337 | 550 | 537 | 719 | 0.80 (0.65-0.98) | 0.04 |
| *Measure of effect modification on the multiplicative scale: Ratio of OR 0.92 (0.69-1.24), p-value 0.60* | | | | | |  |  |
|  |  | **Low social cohesion** | | **High social cohesion** | | Adjusted odds of social cohesion within intervention strata  (aOR; 95% CI) | **p-value** |
| Intervention status | **SHG / non-SHG (n=3,257/1,055)** | non-SHG  (n=1634) | SHG (n=492) | Non-SHG (n=1623) | SHG (n=563) |  |  |
| **Control** |  | 857 | 215 | 840 | 246 | 1.02 (0.79-1.32) | 0.87 |
| **Intervention** |  | 777 | 277 | 783 | 317 | 1.08 (0.88-1.33) | 0.45 |
| *Measure of effect modification on the multiplicative scale: Ratio of OR 1.06 (0.76-1.49), p-value 0.73* | | | | | |  |  |
|  | **EW / non-EW**  **(n=1,835 /2,477)** | EW (n=1006) | non-EW (n=1120) | EW (n=829) | Non-EW (n=1357) |  |  |
| **Control** | 953/1,205 | 524 | 548 | 429 | 657 | 1.41 (1.04-1.89) | 0.025 |
| **Intervention** | 882/1,272 | 482 | 572 | 400 | 700 | 1.31 (0.99-1.73) | 0.055 |
| *Measure of effect modification on the multiplicative scale: Ratio of OR 1.05 (0.73-1.52), p-value 0.79* | | | | | |  |  |
| **adjusted for age, HIV status, sex work duration* | | | | | | | |

| **Table 5: Modification of the effect of self-stigma and social cohesion on quarterly clinic attendance by study arm** | | | | | | | |
| --- | --- | --- | --- | --- | --- | --- | --- |
|  |  | **< Quarterly Clinic Visits (N=2,995)**  n (RDS-II %) | | **≥Quarterly Clinic Visits (N=1,317)**  n (RDS-II %) | | **OR** | **aOR*** |
| **Self-stigma** |  |  |  |  |  |  |  |
| Low | 1,676 | 1109 (37.2) | | 567 (43.2) | | 1 | 1 |
| High | 2,614 | 1869 (62.8) | | 745 (56.8) | | 0.74 (0.62-0.88) | 0.75 (0.62-0.89) |
| **Social cohesion** |  |  |  |  |  |  |  |
| No | 2,126 | 1565 (52.3) | | 561 (42.6) | | 1 | 1 |
| Yes | 2,186 | 1430 (47.8) | | 756 (57.4) | | 1.46 (1.25-1.71) | 1.44 (1.23-1.69) |
| Intervention status |  | High self-stigma | Low self-stigma | High self-stigma | Low self-stigma | Adjusted odds of quarterly clinic visits with low self-stigma within intervention and control sites strata (aOR; 95% CI) | |
| **Control** |  | 1,009 | 550 | 349 | 239 | 0.81 (0.63-1.05) | |
| **Intervention** |  | 860 | 559 | 396 | 328 | 0.72 (0.56-0.92) | |
| *Measure of effect modification on the multiplicative scale: Ratio of OR 0.88 (0.62-1.57), p-value 0.50* | | | | |  |  |  |
| Intervention status |  | No social cohesion | social cohesion | No social cohesion | social cohesion | Adjusted odds of quarterly clinic visits with greater social cohesion within intervention and control sites strata (aOR; 95% CI) | |
| **Control** |  | 826 | 742 | 246 | 344 | 1.56 (1.23-1.97) | |
| **Intervention** |  | 739 | 688 | 315 | 412 | 1.34 (1.08-1.66) | |
| *Measure of effect modification on the multiplicative scale: Ratio of OR 0.86 (0.62-1.18), p-value 0.36* | | | | |  |  |  |

*adjusted for age, HIV status, sex work duration
